# Impact of social distancing measures for preventing coronavirus disease 2019 [COVID-19]: A systematic review and meta-analysis protocol

**DOI:** 10.1101/2020.06.13.20130294

**Authors:** Krishna Regmi, Cho Mar Lwin

## Abstract

**Introduction:** Social distancing measures (SDMs) protect public health from the outbreak of coronavirus disease 2019 (COVID-19). However, the impact of SDMs has been inconsistent and unclear. This study aims to assess the effects of SDMs (e.g. isolation, quarantine) for reducing the transmission of COVID-19.

**Methods and analysis:** We will conduct a systematic review meta-analysis research of both randomised controlled trials and non-randomised controlled trials. We will search MEDLINE, EMBASE, Allied & Complementary Medicine, COVID-19 Research and WHO database on COVID-19 for primary studies assessing the effects of SDMs (e.g. isolation, quarantine) for reducing the transmission of COVID-19, and will be reported in accordance with PRISMA statement. The PRISMA-P checklist will be used while preparing this protocol. We will use Joanna Briggs Institute guidelines (JBI Critical Appraisal Checklists) to assess the methodological qualities and synthesised performing thematic analysis. Two reviewers will independently screen the papers and extracted data. If sufficient data are available, the random-effects model for meta-analysis will be performed to measure the effect size of SDMs or the strengths of relationships. To assess the heterogeneity of effects, I^2^ together with the observed effects (Q-value, with degrees of freedom) will be used to provide the true effects in the analysis.

**Ethics and dissemination:** Ethics approval and consent will not be required for this systematic review of the literature as it does not involve human participation. We will be able to disseminate the study findings using the following strategies: we will be publishing at least one paper in peer-reviewed journals, and an abstract will be presented at suitable national/international conferences or workshops. We will also share important information with public health authorities as well as with the World Health Organization. In addition, we may post the submitted manuscript under review to bioRxiv, medRxiv, or other relevant pre-print servers.

**Strengths and limitations of this study:**

- To our knowledge, this study will be the first systematic review to examine the impact of social distancing measures to reduce transmission of COVID-19.
- This study will offer highest level of evidence for informed decisions, drawing a broader framework.
- This protocol reduces the possibility of duplication, provides transparency to the methods and procedures that will be used, minimise potential biases and allows peer-review.
- This research is not externally funded, and therefore time and resource will be constrained.
- If included studies will be variable in sample size, quality and population, which may open to bias, and the heterogeneity of data will preclude a meaningful meta-analysis to measure the impact of specific SDMs

## Introduction

Coronavirus disease 2019 (COVID-19; caused by severe acute respiratory syndrome coronavirus 2 [SARS-CoV-2]), emerged in Wuhan, China in December 2019, has been the biggest challenge for us in our lifetime posing a global public health threat. At the time of writing (1/6/20) WHO COVID-19 Situation Dashboard reported that this virus has already affected 216 countries with approximately 5,939,234 confirmed cases and 367,255 confirmed deaths; a fatality rate of approximately 6.18%, i.e. more than six deaths in every 100 confirmed cases. The highest number of confirmed cases were reported in the Americas (2,743,793) followed by Europe (2,142,547), Eastern Mediterranean (504,001) and South-East Asia (264, 015), whereas Western Pacific and Africa reported relatively low cases i.e. 182, 527 and 100,610 respectively.^1^ In Europe, the UK has become the ‘epicentre’ of the pandemic.

Based on reported cases and deaths, this disease is portrayed as a great equaliser, but 1:10 reported infections were among health professionals, e.g. medical doctors, nurses and other healthcare professionals. Evidence further indicates that in England, Black, Asian and Minority Ethnic (BAME) groups recorded higher mortality, ranging from 1.5 (in Asian) to 7.3 (in Black Caribbean population) times compared to white individuals.^2^ Similarly, COVID-19 mortality rate in the US for African Americans was 2.4-2.7 times more than white individuals. However, deaths are not consistent across these groups. Several factors could be considered, e.g. ethnicity, age, sex, co-morbidities (diabetes, renal conditions), occupation, socioeconomic status, multifamily and multigenerational households.^2–4^

Similarly, it is difficult to predict an exact future, but recent data from Johns Hopkins University reported that global COVID-19 deaths have surpass over 370,000 worldwide ^5^. Imperial College London highlights that this outbreak could kill 40 million people this year without public health measures (e.g. case finding, contact tracing and testing, and strict quarantine).^6^ Evidence suggests that the number of cases reported would possibly “represent an underestimation of the true burden due to lack of surveillance and diagnostic capacity”^7^ as well as pharmaceuticals to manage severe COVID-19.^8^

Several countries, including the UK, USA and other EU countries are adopting SDMs as a form of non-pharmaceutical or physical intervention. Social distancing is defined as a measure to ban large gatherings and advise individuals not to socialize outside their households by closing borders, some public places, schools and universities; isolation/quarantine, physical distancing and room separation to isolate symptomatic individuals and their contacts; and large-scale lockdowns of populations by staying at least 2m apart aiming to minimize mixing of infectious patients with susceptibles.^8^ WHO recommends case finding, testing, isolation, contact tracing and quarantine of close contacts.^9^

A preliminary scan of the literature demonstrated some research on COVID-19 from China, South Korea, UK, USA and other countries, but these are very limited systemically reviewed or synthesised. Several rapid reviews and summaries have been covered on COVID-19 epidemiology,^10,11^ the effectiveness of real-time PCR for diagnosis,^12^ effects of school closure,^13^ quarantine,^14,15^ social distancing^16^ (whose study was primarily based on two previous reviews^17,18^ on influenza conducted in 2012 and 2018, respectively), and mathematical modelling studies incorporating the effect of social distancing.^8,19–27^ These models would generally help to “predict epidemic curve representing the number of infections caused by the virus over time.”^28^

Recently, few systematic review and meta-analysis conducted to investigate the optimum distance for avoiding transmissions and ethnicity and clinical outcomes.^4,29^ Cochrane further conducted three studies. First, a rapid review in 2020, involving 29 studies on COVID-19, SARS, MERS plus other viruses from China, UK, South Korea and Japan.^30^ Second, a rapid qualitative evidence synthesis conduced in 2020 capturing 36 studies from Asia, Africa, Central and North America and Australia examined healthcare workers’ adherence and enablers or challenges associated with infection control guidelines for respiratory infections. Another study examined 67 studies including RCTs and observational studies exploring the role of physical interventions for reducing the spread of respiratory viruses, and found no evidence regarding screening at entry ports and social distancing.^31^

Lewnard and Lo^7^ and Michigan Medicine Projections^32^ reported that combined SDMs or interventions using social isolation, quarantine, school closure, and workplace distancing appeared effective in reducing COVID-19 compared to no interventions at all. This approach, however, reported considerable challenges, e.g. societal disruption, social isolation/rejection, mental stress and psychological trauma, lack of tests and testing facilities, poor contact tracing, lack of surveillance. None of these studies examined the SDMs factors in reducing the transmission of COVID-19 systematically. We proposed a systematic review to assess the effects of SDMs (e.g. isolation, quarantine) for reducing the transmission of COVID-19.

### Review question

What has been the impact of social distancing measures for preventing coronavirus disease 2019 [COVID-19]?

## Methods and designs

This study will utilise a systematic review (SR), which will consider both randomised controlled trials and non-randomised trials (prospective and retrospective observational studies) of good-quality studies. SR is a research method that reviews relevant research literature, using systematic and explicit, accountable methods, to answer a specific research question objective.^33^ Meta-analysis includes the statistical analysis for combining the results of a number of individual studies to produce summary results, e.g. pooled research studies.^34^ The Preferred Reporting Items for Systematic Reviews and Meta-Analysis Protocols (PRISMA-P) checklist has been used in the preparation of this protocol.^35^

### Criteria for considering studies for review

Inclusion criteria

1. Primary research describing SDMs, e.g. social distance, isolation and quarantine across all age groups.
2. Research reporting different factors and SDMs or social distancing interventions, e.g., social distance by avoiding crowds and restricting movement, isolating ill people and quarantine of exposed people (as a secondary outcome) and reducing transmission of COVID-19 trend (as a primary outcome). Additional outcomes include – anxiety, depressions, physical and psychological distress,
3. Published peer-reviewed article using randomised controlled trials and non-randomised controlled trials
4. Articles published in English language regardless of the location (or settings) of the studies, up to May 2020. [We proposed to collect data from July/August until October 2020 for the study]

Exclusion criteria

1. Articles published in narrative reviews, modelling studies, opinion pieces, letters, news, editorials, perspectives, commentaries and any other publications lacking primary data, including grey literatures.
2. Studies deemed to have overall low quality.

### Search strategy to identify relevant studies

Five major databases will be searched: MEDLINE, EMBASE, Allied & Complementary Medicine, COVID-19 Research and WHO database on COVID-19. The literature search use the following terms: “social distancing measures”, “social distancing”, “quarantine”, “patient isolation” combined with “COVID-19”. Primary search terms are SDMs (all synonyms) and COVID-19 (all synonyms) using ‘Textword searching’ – searching for a word or phrase appearing anywhere in the document, where the document is the citation (article title, journal name, author), not the full text of an article, and ‘Thesaurus (MeSH, EMTREE) searching’, employing Boolean operators and truncations. To maximise sensitivity, a broad search strategy will be designed as shown in table 1. The ‘Related Articles’ including the *best match* and *most recent* features in PubMed will be consulted. Searches will also be supplemented by reviewing the reference lists (‘references of references’) of selected articles to find any other relevant papers. We will also ask subject experts/information specialists from authors’ Universities to verify the research strategy, ensuring its comprehensiveness.

**Table 1.** Search strategy for the MEDLINE

| Search | Search terms will be modified as needed for use in other databases |
| --- | --- |
| #1 | social distancing |
| #2 | social distance |
| #3 | distancing |
| #4 | isolation |
| #5 | patient isolation |
| #6 | patient isolators |
| #7 | physical contact |
| #8 | physical distancing |
| #9 | quarantine |
| #10 | quarantined |
| #11 | #1 OR #2 OR #3 OR #4 OR#5 OR #6 OR #7 OR #8 OR#9 OR #10 |
| #12 | COVID-19 |
| #13 | 2019 ncov |
| #14 | 2019-nCoV |
| #15 | 2019 novel coronavirus |
| #16 | betacoronavirus |
| #17 | Wuhan coronavirus |
| #18 | coronavirus infections |
| #19 | covid 19 pandemic |
| #20 | sars cov 2 |
| #21 | SARS-CoV-2 |
| #22 | sars virus |
| #23 | #12 OR #13 OR #14 OR #15 OR#16 OR #17 OR #18 OR #19 OR#20<br>OR #21 OR#22 |
| #24 | clinical trials |
| #25 | cross-sectional studies |
| #26 | Survey |
| #27 | epidemiologic studies |
| #28 | Quantitative research |
| #29 | #24 OR #25 OR #26 OR #27 OR#28 |

### Selection of studies

The citations identified through the searches will be imported into Mendeley Reference Manager (https://www.mendeley.com/). All studies emerging from the databases will be screened twice: i) screening of screening of titles, abstracts with two reviewer against minimum inclusion criteria, and ii) review of full text. We will use the standard PRISMA flow diagram to provide the process of study selection (figure 2).^36^

**Figure 1.**
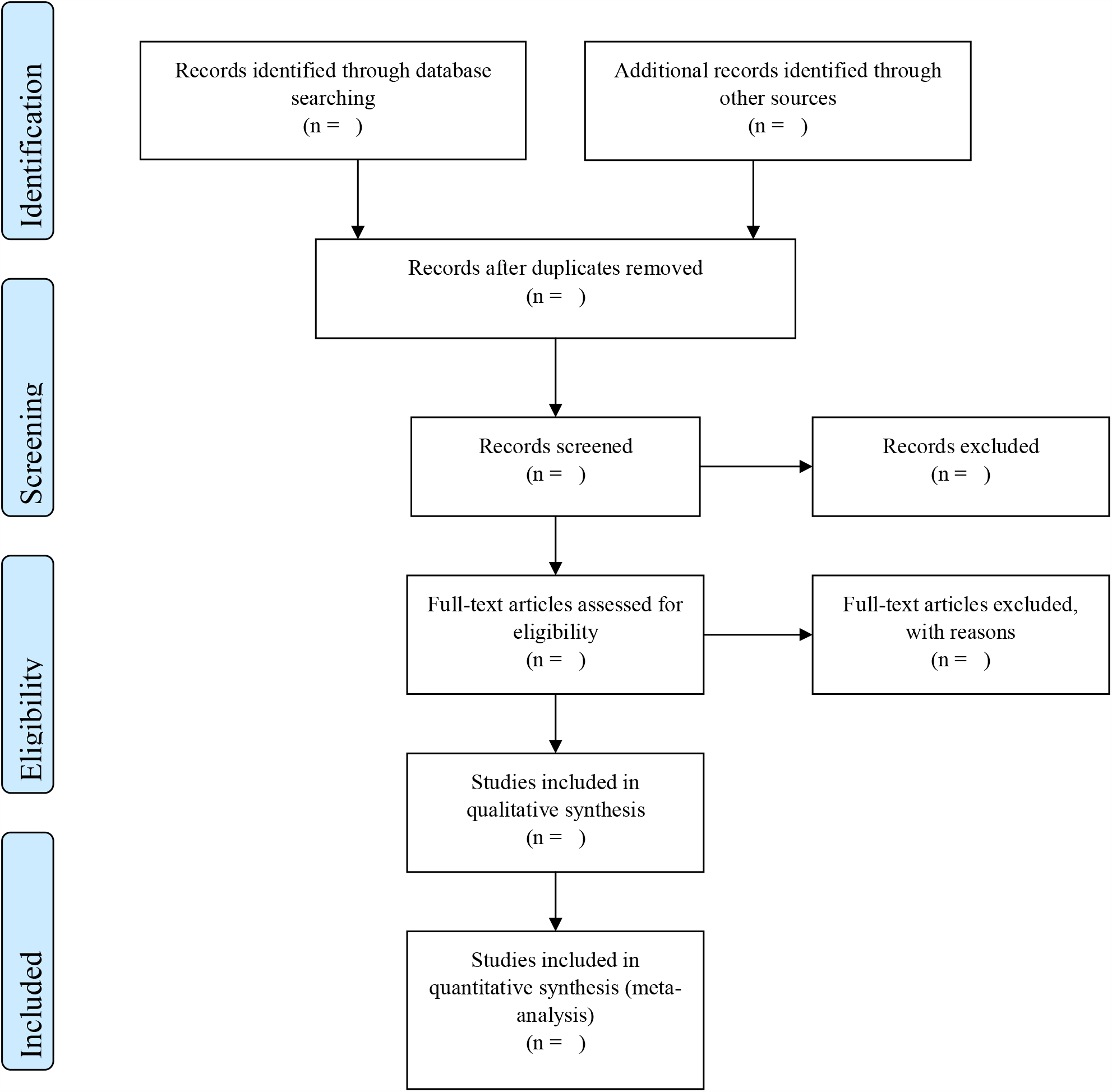
PRISMA flow diagram

### Quality appraisal of included studies

Joanna Briggs Institute (JBI) critical appraisal checklists for randomised controlled trials and non-randomised controlled trials will use to assess the methodological qualities ^37^ (Box 1). All included studies will assess by two reviewers (KR, CML) using the standardised questions 4-item checklists i.e. Yes, No, Unclear and Not Applicable and the results will use to inform synthesis and interpretation of the findings. To facilitate comparison of appraisal processes, all reviewers will record the rationale for inclusion or exclusion, and discrepancies will discuss and resolve by consensus.

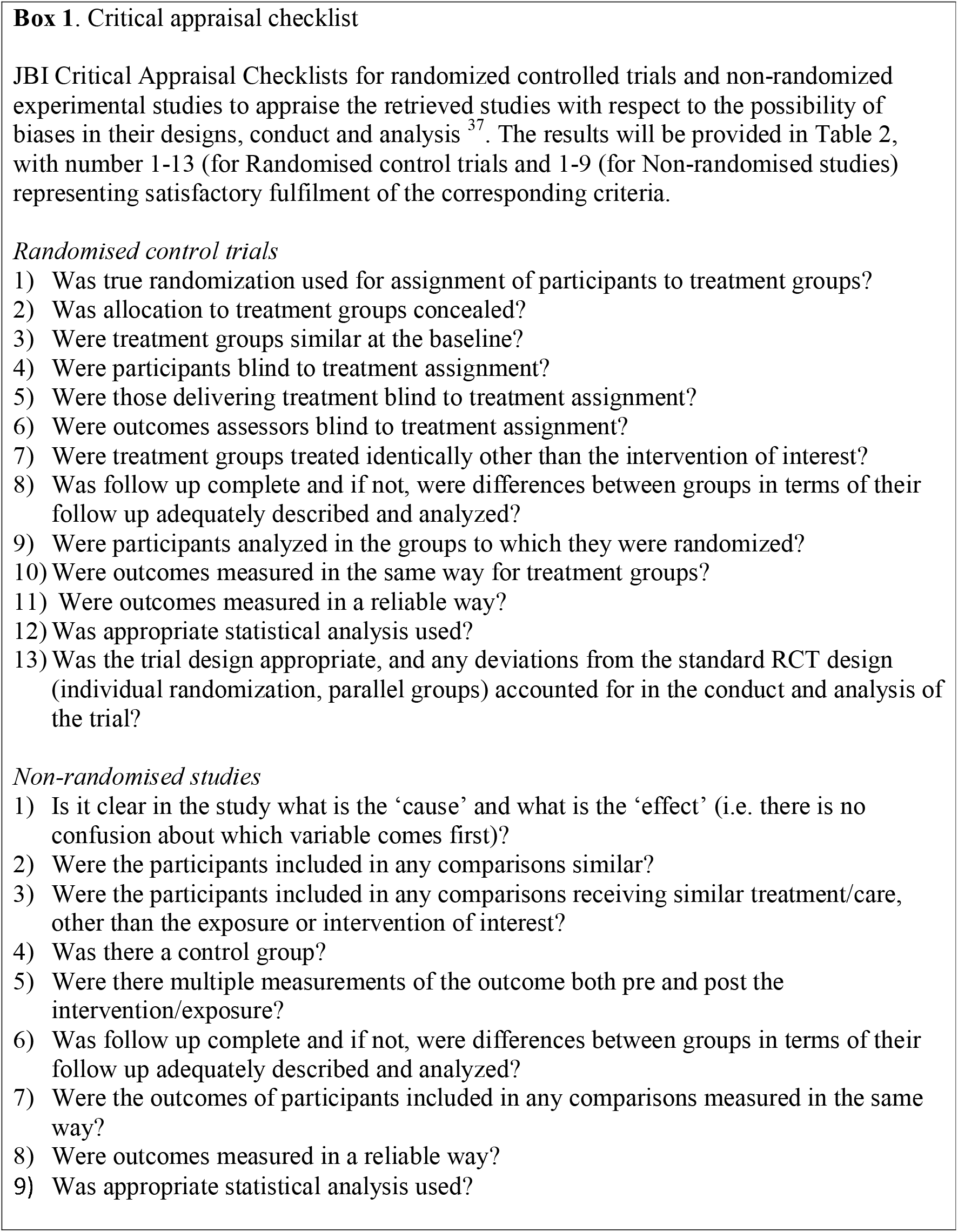

### Assessment of reporting biases

Publication bias, often called reporting bias and dissemination bias, refers to the concern that studies which report relatively large effects are more likely to be published as compared to studies reporting smaller effects.^38^ Similarly, published studies that include multiple outcomes would be more likely to report the outcomes than if they showed statistically significant results.^39^ One approach to address the publication bias is to follow the Trim and Fill procedures, i.e. assessing asymmetry or symmetry in the Funnel plot if more than 10 eligible studies are identified. This approach would estimate the extent of bias or estimate of the adjusted effect size.^40^ We will use this approach while assessing the publication bias in the included studies, but Borenstein^38(p.165)^ warns that the presence of bias will not automatically invalidate the results.

### Data analysis and synthesis

A narrative synthesis, using thematic analysis, will be conducted for the included studies. We will also provide a descriptive numerical summary. We will use risk ratios (RRs), mean differences (MD), or standardised mean differences (SMD, where applicable, will be used for the dichotomous and continuous outcomes respectively.^38^

If sufficient data are available, i.e. identical on important factors and addressing the same fundamental question, to make an inference to a universe of comparable studies, the random-effects model for meta-analysis will be employed for the analysis to measure the effect size of SDMs or the strengths of relationships using the software Comprehensive Meta-Analysis (CMA, version 3. https://www.meta-analysis.com/pages/new_v3.php?cart=BT2P4569026). The purpose of using a random-effects model in the analysis is “to incorporate the assumption that the different studies are estimating different, yet related, intervention effects”.^41^ To assess the heterogeneity of effects, I^2^ together with the observed effects (Q-value, with degrees of freedom) will be used to provide the true effects in the analysis. Q-value is the sum of the squared deviations of all effect sizes from the mean effect size. Generally, this value is on a standardised scale, so that a large deviation gets more weight if the estimate is precise, and less weight if the estimate is imprecise.^42^ In fact, I^2^ statistics does not tell us how much heterogeneity there is, but it tells what proportion of the observed variance reflects in true effect sizes rather than the sampling error. As such, it provides some context for understanding the forest plot.^43^ If I^2^ statistics is low (near zero), then most of the variable in the forest plot is due to sampling error. Conversely, if I^2^ statistics is very high (say, more than 75%) then most of the variance in the forest plot is due to variance in true effects. If we could somehow plot the variance of true effects, most of the variance would remain.^41^

### Tabulating the included studies

Data from eligible studies will be extracted independently by two reviewers based on the summary of review studies (Table 2). As Rodgers and colleagues confirm, this would not only improve the process of transparency by better understanding what sorts of data extracted from which studies, but also recognising the contribution made by each study to the overall synthesis.^44^ In addition, such tables will demonstrate how the individual study area contributes to the reviewers’ final conclusion.

**Table 2.**
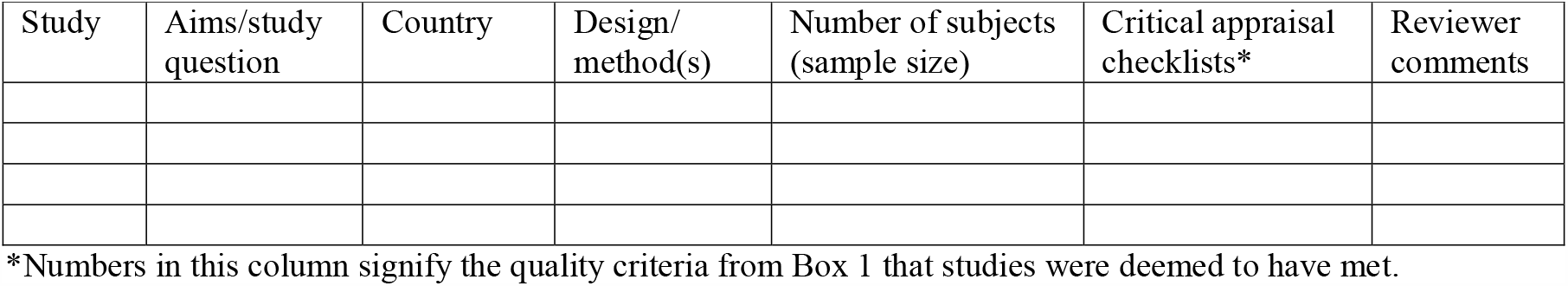
Summary of reviewed studies

### Dealing with missing data

In the case of missing data that might be important to summarise/synthesise the findings of the study or details of the studies are unclear, corresponding authors of included studies will be contacted.

### Sub-group analysis

An a priori sub-group analysis will be planned, if data available, for:

a. social distancing;
b. isolation; and
c. quarantine.

### Risk of bias

Risk of bias will be examined, as it provides the variation, e.g. heterogeneity in the results of the studies included in the study. As Higgins et al.^41^ argue, rigorously conducted studies in the systematic review would provide more truthful results, and the results from the studies of variable validity would give either false negative or false positive conclusions. Therefore, assessing the risk of bias in all studies in any review is important. In assessing risk, we will create a table with a row for every relevant type of potential bias, and then classify each study on each row as having a low, unclear, or high risk of bias. In this study, the issue of bias will be kept separate from the core analysis – meaning analysis will be performed without worrying about the quality/bias. We will then use the risk of bias table to provide the context for the analysis.^38^ As Borenstein^38(p.326)^ suggests, “if the analysis shows a clinically and/or substantially important effect, we will assess the entirety of the evidence by considering the risk of bias as well.” Generally, the bias table provides the type of bias (e.g., selective reporting of outcomes, random sequence generation, allocation of concealment, blinding of participants, personnel and assessors, incomplete outcome data and other potential threats to validity) in each study. If, for example, most rows are unshaded then that it is considered a low risk of bias, whereas if some (or all) rows are either partly shaded or dark (risk of bias will be either unclear or high), this would provide relatively less confidence in the results.^41^ We will use both RoB 2 tool ^45^ for randomised and ROBINS-I tool ^46^ for non-randomised trials while assessing the risk of bias.

### Patient and public involvement

As this is a protocol for a systematic review and meta-analysis, neither patients nor public participation will be directly involved, and ethics approval and consent will not be required either.

### Dissemination

We will be able to disseminate the study findings using the following strategies: we will be publishing at least one paper in peer-reviewed journals, and an abstract will be presented at suitable national/international conferences or workshops. We will also share important information with public health authorities as well as with the World Health Organization. In addition, we may post the submitted manuscript under review to bioRxiv, medRxiv, or other relevant pre-print servers.

## Discussion

To our knowledge, this study will be the first systematic review to examine the effects of SDMs (e.g. isolation, quarantine) for reducing the transmission of COVID-19. enablers and barriers impacting SDMs to reduce transmission of COVID-19. Social distancing becomes a highly charged topic creating a lieu of debate among the politicians, economists, medical and public health professions. The likelihood is that COVID-19 will become endemic, which suggests long-term behavioural adjustments.^47^ Similarly, we argued that social distancing is not part of the culture in either developed or developing countries, for different reasons.^48^ In developing countries, it is more related to population density, crowding, workplace conditions etc., such as overcrowding in public transport. In developed countries such as Switzerland, people were still following Swiss kiss as late as 20 March, when COVID-19 was already peaking. Similarly, some evidence shows some relationships between social distancing and economic aspects: poverty, living in slums etc. in developing countries; marginalized populations in developed countries. A similar issue has also been reported in the previous study.^49^ Therefore, there is a need to completely change the way the economy, businesses, and life are organised to protect the vulnerable groups such as homeless, disabled, undocumented migrant workers and inmates. Similarly, home life should be looked at, as evidence suggests we need to change the way we interact at home, for example, with vulnerable family members – elderly, pregnant, immunocompromised due to chronic disease or protracted illnesses, at least until the pandemic is over, e.g. curbing the possibility of transferring the disease to the elderly. A recent descriptive review of data on disparities in the risk and outcomes from COVID19 in the UK has reported that:

> “The largest disparity found was by age. Among people already diagnosed with COVID19, people who were 80 or older were seventy times more likely to die than those under 40. Risk of dying among those diagnosed with COVID-19 was also higher in males than females; higher in those living in the more deprived areas than those living in the least deprived; and higher in those in Black, Asian and Minority Ethnic (BAME) groups than in White ethnic groups” ^3^

Marmot et al.^50(p.13)^ also argued that: “There are clear socioeconomic gradients in preventable mortality. The poorest areas have the highest preventable mortality rates and the richest areas have the lowest.” We argue that public health has failed to convince politicians to take rapid action on prevention of spread or prepare for necessary treatment arrangements. Several authors reported that the “structure and capacity of our depleted healthcare system are now largely driving the response to this epidemic” and most likely “it will continue to do so until services that support local communicable disease control are rebuilt and reintegrated.”^51,52^

The potential limitations of this study would be that if the retrieved studies would be variable in sample size, quality and population, which may open to bias, and the heterogeneity of data precludes a meaningful meta-analysis to measure the impact of specific SDMs for COVID-19, therefore the findings might warrant generalisation. Second, methodologies might be poorly reported (mostly due to preprints - postings in MedRxiv), lacking comprehensive strategies for sampling and procedures, and lacking detail in data gathering and analysis. Wolkewitz and Puljak^53^ warned that: “there are many methodological challenges related to producing, gathering, analysing, reporting and publishing data in condensed timelines required during a pandemic.” Third, searching “social distancing” in different databases might be challenging mainly due to rapidly-growing COVID-19 studies in PubMed and other search interfaces, which are not visible in the major search databases (PubMed, EMBASE) due to i) indexing, and ii) often bibliographic databases failed to capture preprint and unpublished studies including registered clinical trials,^54,55^ and the majority are commentaries, news, perspectives or opinions.^53^ Finally, this research is not externally funded, and therefore time and resource will be constrained.

Nevertheless, this study will add to the literature on highlighting the major enablers and barriers of SDM in controlling COVID-19 in public health policy and interventions: i) given the fact that there is no vaccine or treatment available at the time of writing, and ii) there have been limited robust published studies of SDM success factors, with most studies exploring the process rather than hard or tangible outcomes. In addition, this review will provide a basis for developing the best methods and approaches in terms of developing objective measures and interventions to establish the link between different factors and SDMs (as a secondary outcome) and reducing transmission of COVID-19 trend (as a primary outcome) effectively, efficiently and equitably.

## Data Availability

NA

## Contributors

KR conceived and designed the research with the advice from CML; KR wrote the first draft;

KR and CML reviewed and contributed to drafting, revising and finalising the manuscript.

All authors have reviewed and approved the final version of the manuscript and have given their permission for publication.

## Funding statement

The authors received no financial support for this research study.

## Competing interests

The authors declare that they have no competing interests.

## Patient consent for publication

Not required

## Notes

### Competing Interest Statement

The authors have declared no competing interest.

### Author Declarations

Ethical approval - Not applicable

